# Approximation of bolus arrival time by integrating MR arterial spin labeling and SPECT: theory and clinical application

**DOI:** 10.1101/2023.07.10.23292486

**Authors:** Kazumasa Otomo, Shinichiro Sugiyama, Hideki Ota, Hiroyuki Sakata, Hidenori Endo

## Abstract

**Background:** Magnetic resonance arterial spin labeling (ASL) imaging with multiple post-labeling delays (PLDs) provides the bolus arrival time (BAT) as well as cerebral blood flow (CBF) to characterize the cerebral hemodynamics. However, the complexity of data acquisition and processing inhibits the calculation of BAT. We developed a simple method for approximating BAT using single-PLD ASL imaging and single-photon emission computed tomography (SPECT). We conducted a proof-of-concept study in patients with carotid artery stenosis.

**Methods:** We introduced the ASL/SPECT ratio, calculated by dividing the tissue magnetization in pulsed continuous ASL by the CBF measured using SPECT. In theory, the ASL/SPECT ratio has a positive relationship with BAT. Our proof-of-concept study included 63 patients who underwent carotid endarterectomy (CEA) in our hospital from 2014 to 2019. After preprocessing the ASL and SPECT datasets using three-dimensional stereotactic surface projection, we calculated the ASL/SPECT ratio at each voxel. We investigated the correlation between the preoperative BAT and the postoperative CBF.

**Results:** We found a positive correlation between the delay of BAT and the increase rate of CBF in the ipsilateral middle cerebral artery territory (Pearson’s correlation coefficient, 0.444; 95% confidence interval, 0.220–0.623; p=0.000269). Four patients (6.3%) presented with hyper-perfusion phenomenon. Visualization of the BAT revealed that the area prone to postoperative hyper-perfusion presented with a delayed preoperative BAT.

**Conclusions:** Our findings suggest the feasibility of the BAT approximated using ASL and SPECT in patients with chronic steno-occlusive cerebrovascular diseases. The proposed concept is also applicable to ASL and any modalities that measure CBF.

## Introduction

Arterial spin labeling (ASL) is a noninvasive magnetic resonance (MR) technique used to examine cerebral hemodynamics (1). ASL datasets obtained at multiple post-labeling delays (PLDs) enabled us to assess not only cerebral blood flow (CBF) but also bolus arrival time (BAT) (2-4). The BAT is defined as the time required for labeled spins to reach the target tissues. Several previous studies reported the ability of BAT to characterize ischemic status in patients with steno-occlusive cerebrovascular diseases, such as acute stroke, chronic internal carotid artery (ICA) occlusion, or Moyamoya disease (4–7). However, the complexity of data processing accompanied with multi-PLD ASL imaging has reduced the feasibility of BAT in the neuroradiological field.

We developed a simple method for approximating BAT by integrating a single-PLD ASL imaging and other medical modalities that measure cerebral blood flow (CBF). In this study, we used single-photon emission computed tomography (SPECT) for measuring the CBF. We propose a theory in which we convert the difference in ASL signal intensity between two regions of interest (ROIs) into a difference in BAT by mathematical integration of ASL and SPECT datasets. In addition, we conducted a proof-of-concept study, in which we applied the proposed method to the hemodynamic assessment of patients with ICA stenosis.

## Methods

### Introduction of the ASL/SPECT ratio

Based on the single-compartment theory, the difference in tissue magnetization (*ΔM*(*t*)) between the control and labeled images in three-dimensional (3D) pulsed continuous ASL (pCASL) imaging is described using Eqs. (1)–(3) (8-11):

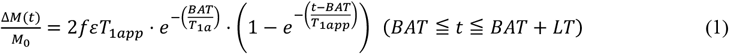

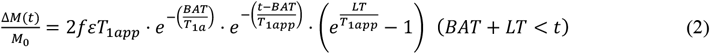

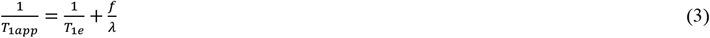

where *f* is blood flow, *ε* is inversion efficacy of labeling pulse, *BAT* is bolus arrival time from labeling plane to image slice, *LT* is labeling time, *T*_l*a*_ is longitudinal relaxation time of water in blood, *T*_l*e*_ is longitudinal relaxation time of water in tissue, and λ is tissue blood partition coefficient of water. A fully relaxed blood spin (*M*_0_) is typically used for the local signal intensity of proton density images. In Figure 1, we demonstrate the tissue magnetization (*ΔM*(*t*)) with time after the start of labeling based on Eqs. (1)–(3).

**Figure 1.**
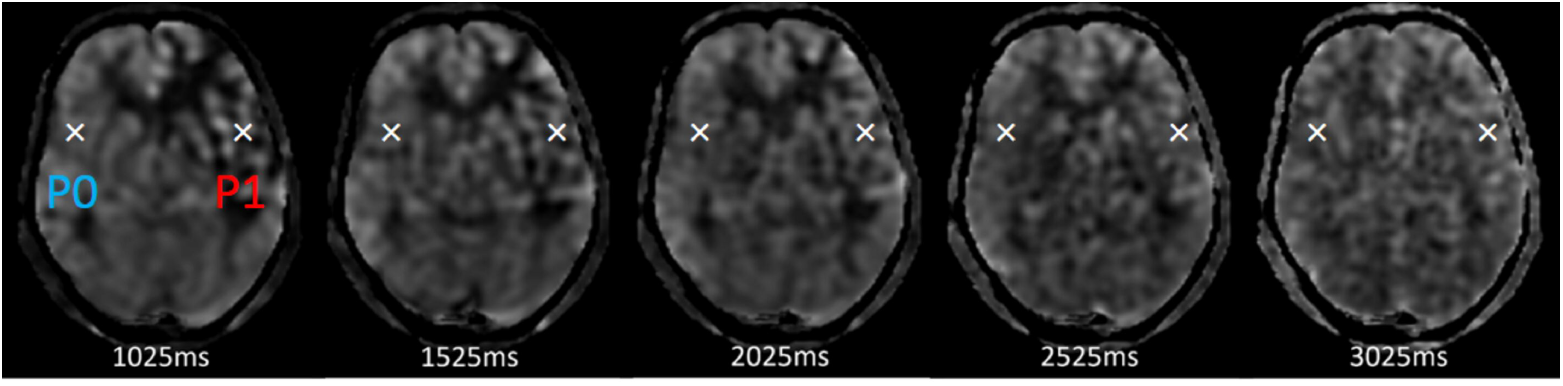
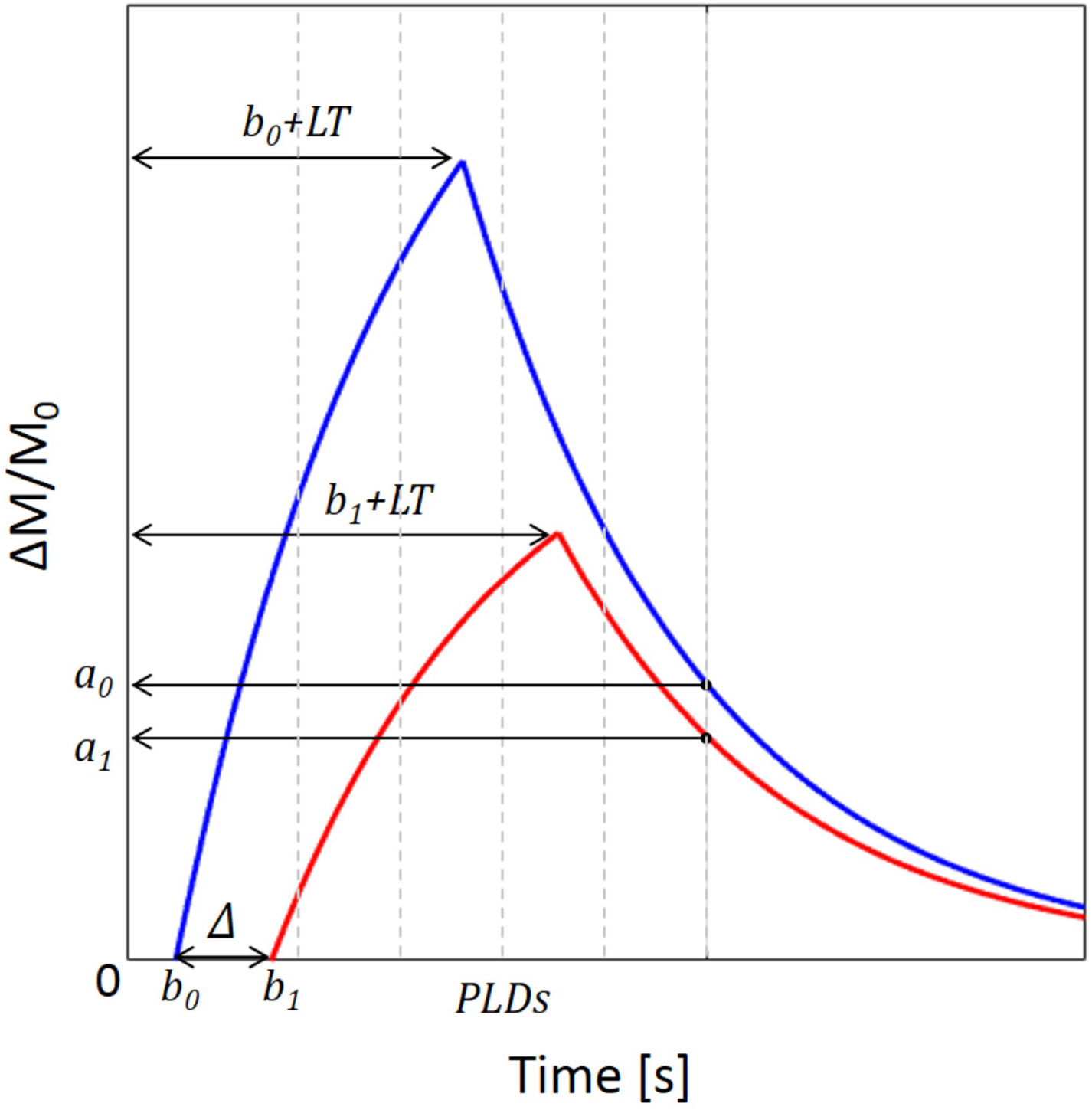
Schemas showing the concept of the ASL/SPECT ratio. (**A**) ASL images obtained at multiple PLDs (1.025–3.025 (s)) in a patient with left carotid artery stenosis. Note the asymmetry in signal intensity derived from the difference in CBF and BAT. Conversely, the temporal variation of the signal intensity under the constant CBF enables the approximation of BAT using multi-PLD ASL imaging. We compared SPECT and single-PLD ASL data to approximate BAT. (**B**) Schematic graphs showing tissue magnetization in pCASL imaging. Consider two points: P_1_ at the ipsilateral side (red) and P_0_ at the contralateral side (blue) shown in Figure 1A. The exponential curves show the tissue magnetization (ΔM/M_0_) with time after the start of labeling (t = 0) at P_0_ (blue line) and P_1_ (red line), based on a single-compartment model. The tissue magnetization at a certain PLD is assumed to be a_0_ at P_0_ and a_1_ at P_1_, and the BAT at P_0_ and P_1_ is b_0_ and b_1_, respectively. The subtraction of b_0_ from b_1_ leaves the delay of BAT at P_1_ (Δ (s)). When SPECT provides CBF at P_0_ and P_1_ as c_0_ and c_1_, we can calculate the ASL/SPECT ratios as a_0_/c_0_ at P_0_ and a_1_/c_1_ at P_1,_ respectively. Under the condition that the PLD is longer than the sum of BAT and LT, we can approximate Δ from the logarithmic conversion of the ASL/SPECT ratios at P_0_ and P_1._ BAT: bolus arrival time CBF: cerebral blood flow LT: labeling time pCASL: pulsed continuous arterial spin labeling PLD: post-labeling delay SPECT: single-photon emission computed tomography

Let *P*_*k*_(*a*_*k*_, *b*_*k*_, *c*_*k*_) mean that a certain point *P*_*k*_ (*k* = 0, 1, 2, ⋯, *N*) in brain tissue has Δ*M*(*t*)/*M*_0_= *a*_*k*_ and *BAT* = *b*_*k*_ in the pCASL imaging, and *f* = *c*_*k*_ in SPECT. Here, we define the ASL/SPECT ratio (*γ*_*k*_) using Eq. (4):

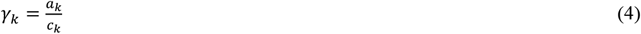

Now we consider two points: *P*_*k*_(*a*_*k*_, *b*_*k*_, *c*_*k*_) and *P*_*l*_(*a*_*l*_, *b*_*l*_, *c*_*l*_) (*k, l* = 1, 2, ⋯, *N*).

The difference in the BAT between the two points (Δ) is obtained using Eq. (5):

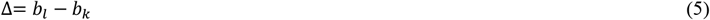

Under the condition that *t* = *PLD* > *BAT* + *LT*, we obtained Eqs.(6) and (7) using Eq. (2):

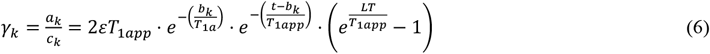

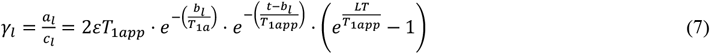

By dividing Eq. (7) by Eq. (6), we get Eq. (8):

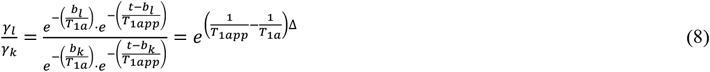

Finally, we obtain Δ using Eqs. (9)–(11):

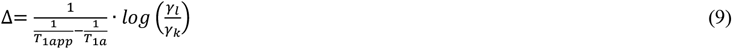

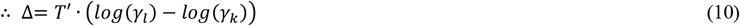

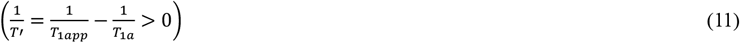

According to Eq. (10), Δ is in proportion to the difference of the logarithmic conversion of the ASL/SPECT ratios. Considering that the logarithmic function increases monotonically, the ASL/SPECT ratio and BAT have a positive relationship.

### Delta ratio for visualization

According to the theorems we have discussed above, we can define the difference in BAT (Δ_*n*_) between a certain reference point (*P*_0_(*a*_0_, *b*_0_, *c*_0_)) and any target points (*P*_*n*_(*a*_*n*_, *b*_*n*_, *c*_*n*_) (*n* = 1, 2, ⋯, *N*)).

Here, we can define the delta ratio (δ_*n*_) using Eq. (12):

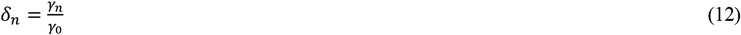

Using Eqs. (9) and (11), we get Eq. (13):

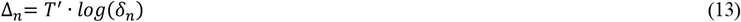

Δ_*n*_ monotonically increases as δ_*n*_ increases. When applying the proposed concept in clinical practice, δ_*n*_ is convenient for clinical visualization because of the simplicity of its calculation without logarithmic conversion.

### Proof-of-concept study

This retrospective study was approved by the Institutional Ethics Review Board of our hospital (2021-0520-4). The requirement for obtaining written informed consent was waived owing to the retrospective nature of the study. This study included all consecutive patients with ICA stenosis who were treated with carotid endarterectomy (CEA) from 2014 to 2019. Patients with major ipsilateral infarction and/or > 50% contralateral ICA stenosis were excluded from the study. The data that support the findings of this study are available from the corresponding author upon request. This study adhered to the STROBE reporting guideline.

### ASL imaging

MR imaging was performed before CEA using a 1.5T MR scanner (Signa HDxt; GE Healthcare, Tokyo, Japan). We acquired 3D pulsed continuous ASL (pCASL) images using the following parameters: repetition time, 4546 [ms]; echo time, 10.5 [ms]; field of view, 24.0 [cm]; number of sampling points, 512; number of sections, 30; number of excitations, 2; bandwidth, 62.50 [Hz]; labeling time, 1.5 [s]. The datasets were obtained at a PLD of 2,525 [ms].

### SPECT

All patients underwent SPECT examinations of ^123^I-iodoamphetamine autoradiography using Infinia Hawkeye 4 (GE Healthcare, Tokyo, Japan) to monitor pre- and postoperative CBF.

### Preprocessing

We preprocessed the 3D-pCASL and SPECT datasets using the 3D stereotactic surface projection method (FALCON; Nihon-Mediphysics, Tokyo, Japan), which fixes position alignment and sets ROIs based on vascular territories (12).

### Approximation of BAT

First, we calculated the ASL/SPECT ratio at each voxel (*γ*_*n*_ (*n* = 1, 2, ⋯, *N*)) using the preprocessed datasets of 3D-pCASL and SPECT. Second, we calculated the average of the ASL/SPECT ratios in ipsilateral and contralateral middle cerebral artery (MCA) territories (*γ* and *γ*_*e*_). Third, we calculated the delta ratio at each voxel (δ_*n*_(*n* = 1, 2, ⋯, *N*)) by dividing *γ*_*n*_ by *γ*_*e*_ and created preoperative maps of the delta ratio (Figure 2). Finally, we approximated the delay in the BAT (Δ) of the ipsilateral MCA territory compared with the contralateral one using Eqs. (9) and (11). We set T1a and T1app to 1.65 [s] and 1.2 [s], respectively (13,14).

**Figure 2.**
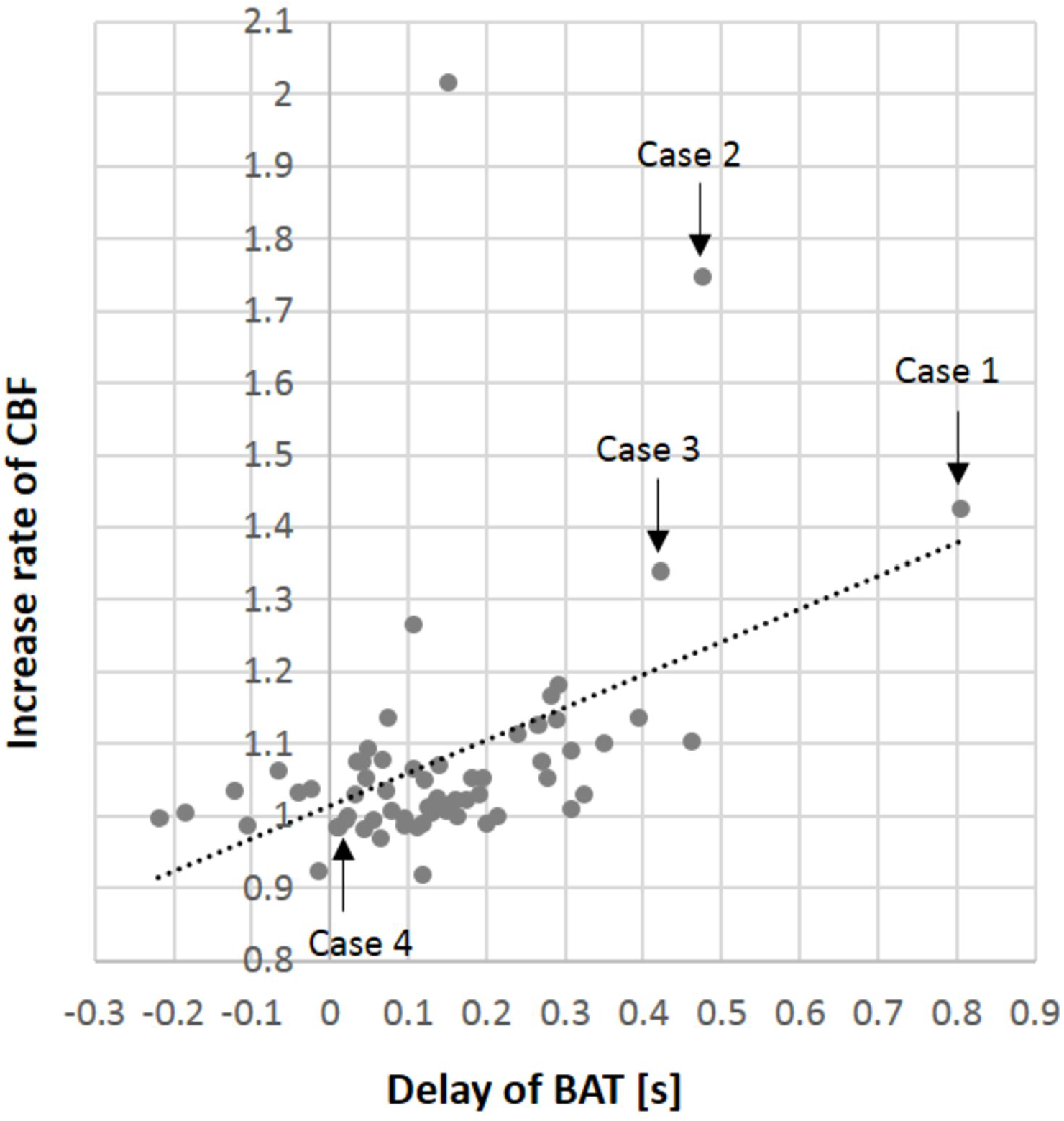
Preoperative BAT delay and postoperative CBF increase The graph shows a positive correlation between the preoperative delay of BAT and the postoperative increase of CBF in the ipsilateral MCA territory. We identified three cases with prolongation of the BAT delay over 0.4 [s] and an increased rate of CBF over 1.25 (cases 1-3). (see also Figure 3)

### Correlation between preoperative and postoperative hemodynamics

We investigated the influence of the preoperative hemodynamic status characterized by BAT on postoperative hemodynamics. We calculated the increased rate of CBF in ipsilateral MCA territory by comparing pre- and postoperative SPECT. We investigated the correlation between the delay of BAT and the increased rate of CBF by regression analysis.

### Statistical analysis

The Mann–Whitney U test was used for parametric statistical analysis. Categorical variables were analyzed using Fisher’s exact test. The linear relationship between continuous variables was estimated using Pearson’s correlation coefficients. Statistical significance was set at p<0.05. All statistical analyses were performed using the open-source statistical packages R and R commander (versions 4.0.3 and 2.7.1, respectively; R Foundation for Statistical Computing, Vienna, Austria).

## Results

This study included 63 patients with carotid artery stenosis treated with CEA. The patients’ mean age was 69.7 years (standard deviation [SD], 6.3). Seven (14.3%) patients were female. The average of the ASL/SPECT ratios in the contralateral and ipsilateral MCA territories were 2.13 (SD, 0.42) and 2.30 (SD, 0.40), respectively. The average of the BAT delay (Δ) in the ipsilateral MCA territory compared with the contralateral one was 0.144 [s] (SD, 0.167).

### BAT delay and CBF increase

Figure 2 shows the influence of the preoperative hemodynamic status characterized by BAT on postoperative CBF in the ipsilateral MCA territory. We found the positive correlation between the delay of BAT and the increased rate of CBF (Pearson’s correlation coefficient, 0.444; 95% confidence interval, 0.220–0.623; p=0.000269).

The mean increased rate of CBF in the ipsilateral MCA territory after CEA was 1.08 (SD, 0.17). We considered that an increase of CBF over 1.25 times (mean plus SD) after CEA represented pathologic hyper-perfusion phenomenon. Four patients (6.3%) presented with hyper-perfusion phenomenon.

### Visualization using delta ratio

In four patients who presented with pathologic hyper-perfusion phenomenon, the delta ratio map indicated the region prone to the hyper-perfusion (Figure 3). On the other hand, the preoperative delta map was symmetrical in cases without a postoperative increase in CBF.

**Figure 3.**
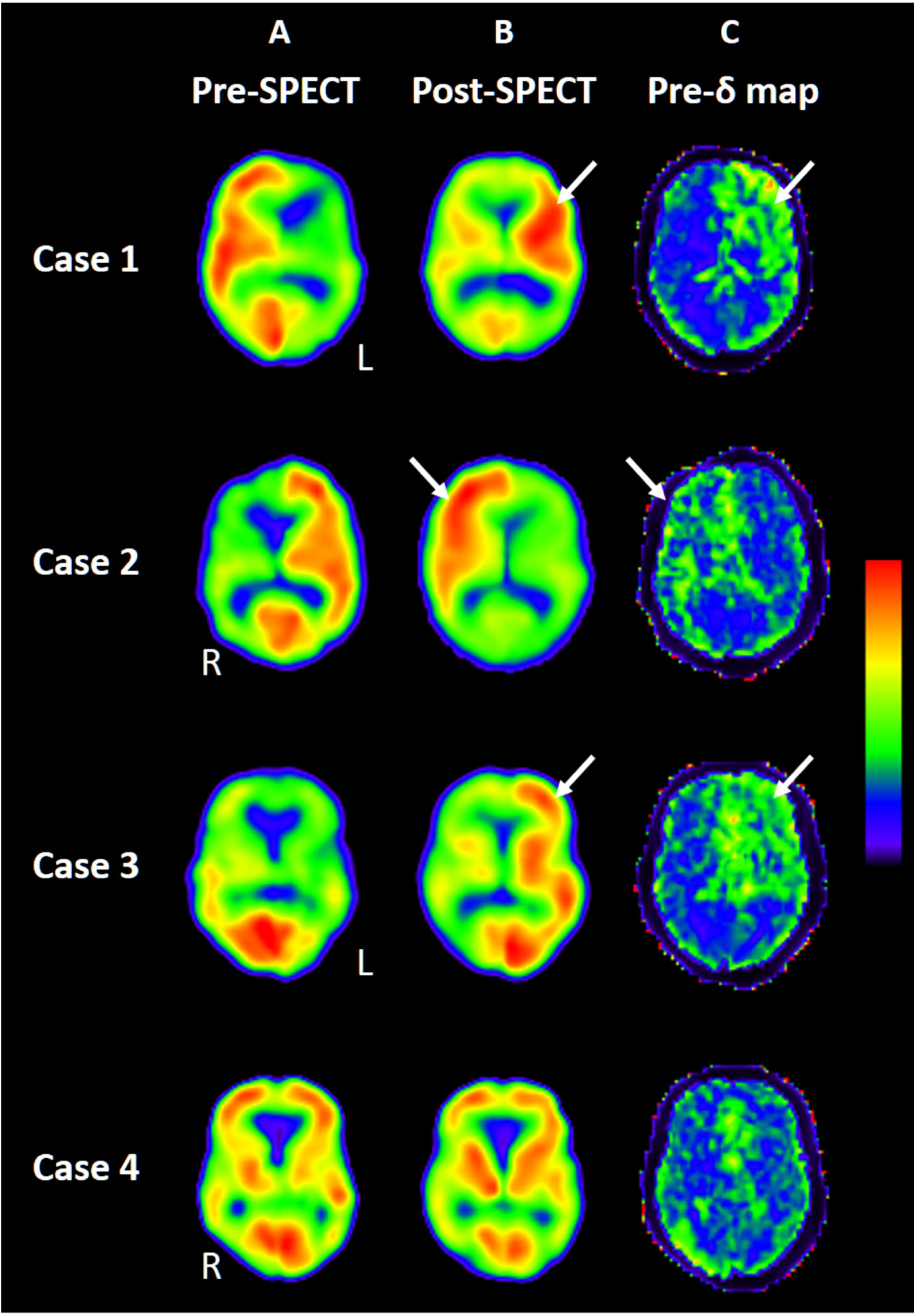
Visualization using delta ratio (**A**) Preoperative SPECT. Affected side is also shown by L or R for left or right, respectively. (**B**) Postoperative SPECT (**C**) Preoperative delta map. Note that the area with postoperative hyper-perfusion phenomenon presents with a high delta ratio (case 1-3; white arrows). The preoperative delta map was symmetrical in a case without postoperative hyper-perfusion phenomenon (case 4). The range of the color bar is as follows: from 15 to 45 (mL/100 mg/min) for SPECT and from 0 to 3.5 for the delta map.

## Discussion

This study proposes a concept for approximating the BAT by integrating ASL and SPECT datasets. Theoretically, the difference in the logarithmic conversion of the ASL/SPECT ratios between the two ROIs is proportional to the difference in BAT. Simply put, the principle depends on the difference in the pharmacokinetics of the tracers in ASL and SPECT. The tracers in ASL are non-accumulative intra-arterial protons, whereas those in SPECT are accumulative radioisotopes generated via venous injection. The former enables the dynamic measurement of CBF for several seconds, whereas the latter provides a static measurement of CBF. In other words, tissue magnetization in ASL is dependent on both CBF and BAT, whereas the gamma rays in SPECT are independent of BAT. Therefore, BAT can be approximated by substituting the CBF measured using SPECT in the equations describing the correlation between CBF and BAT in ASL. In addition to ASL and SPECT, this concept may be applicable to ASL and other modalities that measure CBF.

According to this theoretical proposition, we applied this concept in clinical practice. In patients with ICA stenosis, we calculated the delay of BAT in the ipsilateral MCA territory. In comparison of pre- and postoperative hemodynamics, we found a positive correlation between the preoperative BAT delay and the postoperative CBF increase. By visualizing the delta ratio, which has a positive correlation with BAT delay, we confirmed that the area with a high delta ratio coincided with the area with a high increase in CBF. The prolongation of BAT implies the dissipation of the kinetic energy of blood flow at the stenotic part and a decrease in both blood velocity and cerebral perfusion pressure in the downstream vessels. The decrease of cerebral perfusion pressure causes post-CEA hyper-perfusion syndrome (15). Thus, the results of this study suggest the ability of BAT to characterize hemodynamic impairment in patients with ICA stenosis.

### Limitations

This study had some limitations. The proposed concept requires further validation; the measurement error should be evaluated by comparing the data acquired by conventional multi-PLD ASL. We consider that the BAT obtained using our proposed method might remain a matter of approximation rather than a realistic measurement.

The simplicity of the proposed method might increase the feasibility of approximating the BAT in patients with various types of steno-occlusive cerebrovascular diseases. However, in patients with chronic cerebral ischemia, collateral blood supply frequently develops, and the BAT is remarkably prolonged. In such cases, the single-compartment theory is not applicable to describe tissue magnetization in 3D-pCASL imaging. Further research is required to determine the ASL/SPECT ratio in patients with remarkable prolongation of the BAT.

## Conclusion

This study proposed a simple method for approximating BAT by integrating MR-ASL and SPECT. The results of this study demonstrate the feasibility of BAT, approximated using the proposed method, to characterize ischemic status in patients with chronic steno-occlusive cerebrovascular diseases. The proposed concept is also applicable to ASL and any modalities that measure CBF.

## Data Availability

The data that support the findings of this study are available from the corresponding author upon reasonable request.

## Non-standard Abbreviations and Acronyms

ASL: arterial spin labeling
BAT: bolus arrival time
CBF: cerebral blood flow
CEA: carotid endarterectomy
ICA: internal carotid artery
MCA: middle cerebral artery
MR: magnetic resonance
pCASL: pulsed continuous ASL
PLD: post-labeling delay
ROI: region of interest
SPECT: single-photon emission computed tomography

## Acknowledgments

We thank Dr. Nitin Shivappa for professional English language editing.

